## Supplementary file 1 for "Association between *APOL1* risk variants and the occurrence of sepsis in Black patients hospitalized with infections: a retrospective cohort study"

**Supplementary File 1a. ICD-9-CM and ICD-10-CM codes for infection categories.**

**Supplementary File 1b. ICD-9-CM and ICD-10-CM codes for comorbidities.**

**Supplementary File 1c. List of codes to identify severe renal diseases.**

**Supplementary File 1d. Associations between *APOL1* high-risk genotypes and sepsis-related outcomes.**

**Supplementary File 1e. Demographic summary of patients in the PheWAS analysis.**

**Supplementary File 1a. ICD-9-CM and ICD-10-CM codes for infection categories.**

| Enhanced REGARDS categories(1) | ICD-9-CM | ICD-10-CM |
|---|---|---|
| Intestinal Infectious Diseases | 001.0, 001.1, 001.9, 002.0, 002.1, 002.2, 002.3, 002.9, 003.0, 003.1, 003.20, 003.21, 003.22, 003.23, 003.24, 003.29, 003.8, 003.9, 004.0, 004.1, 004.2, 004.3, 004.8, 004.9, 008.00, 008.01, 008.02, 008.03, 008.04, 008.09, 008.1, 008.2, 008.3, 008.41, 008.42, 008.43, 008.44, 008.45, 008.46, 008.47, 008.49, 008.5, 008.8, 009.0, 009.1, 009.2, 009.3 | A00.0, A00.1, A00.9, A01.00, A01.1, A01.2, A01.3, A01.4, A02.0, A02.1, A02.20, A02.21, A02.22, A02.23, A02.24, A02.29, A02.8, A02.9, A03.0, A03.1, A03.2, A03.3, A03.8, A03.9, A04.4, A04.0, A04.1, A04.2, A04.3, A04.8, A04.5, A04.6, A04.7, A04.9, A08.8, A09 |
| Zoonotic Bacterial Diseases* | 023.0, 023.1, 023.2, 023.3, 023.8, 023.9, 027.0, 027.1, 027.2, 027.8, 027.9 | A23.0, A23.1, A23.2, A23.3, A23.8, A23.9, A32.7, A32.12, A32.9, A32.81, A32.11, A32.89, A26.7, A26.8, A26.9, A28.0, A28.8, A28.9 |
| Other Bacterial Diseases | 032.0, 032.1, 032.2, 032.3, 032.81, 032.82, 032.83, 032.84, 032.85, 032.89, 032.9, 033.0, 033.1, 033.8, 033.9, 034.0, 034.1, 035, 036.0, 036.1, 036.2, 036.3, 036.40, 036.41, 036.42, 036.43, 036.81, 036.82, 036.89, 036.9, 038.0, 038.10, 038.11, 038.12, 038.19, 038.2, 038.3, 038.40, 038.41, 038.42, 038.43, 038.44, 038.49, 038.8, 038.9, 040.0, 040.1, 040.2, 040.3, 040.41, 040.42, 040.81, 040.82, 040.89, 041.00, 041.01, 041.02, 041.03, 041.04, 041.05, 041.09, 041.10, 041.11, 041.12, 041.19, 041.2, 041.3, 041.4, 041.41, 041.42, 041.43, 041.49, 041.5, 041.6, 041.7, 041.81, 041.82, 041.83, 041.84, 041.85, 041.86, 041.89, 041.9 | A36.0, A36.1, A36.89, A36.2, A36.86, A36.81, A36.85, A36.3, A36.83, A36.82, A36.84, A36.9, A37.00, A37.10, A37.80, A37.90, J03.00, J02.0, A38.9, A46, A39.0, A39.81, A39.4, A39.1, A39.50, A39.53, A39.51, A39.52, A39.82, A39.83, A39.89, A39.9, A40.9, A41.2, A41.0, A41.01, Z16, A41.02, A41.1, A41.89, A40.3, A41.4, A41.50, A41.3, A41.51, A41.52, A41.53, A41.59, A41.9, A48.0, A48.8, K90.81, A48.51, A48.52, M60.009, A48.3, B95.5, B95.0, B95.1, B95.4, B95.2, B95.8, B95.61, B95.6, B95.62, B95.7, B95.3, B96.1, B96.2, B96.21, B96.22, B96.23, B96.29, B96.20, B96.3, B96.4, B96.5, B96.0, A49.3, B96.6, B96.7, B96.89, B96.81 |
| Neurologic | 320.0, 320.1, 320.2, 320.3, 320.7, 320.81, 320.82, 320.89, 320.9, 322.0, 322.1, 322.2, 322.9, 324.0, 324.1, 324.9 | G00.0, G00.1, G00.2, G00.3, G01, G00.8, G00.9, G04.2, G03.0, G03.8, G03.1, G03.9, G06.0, G06.1, G06.2 |
| Circulatory | 420.0, 420.90, 420.91, 420.99, 421.0, 421.1, 421.9 | I32, I30.9, I30.0, I30.8, I33.0, I39, I33.9 |
| Respiratory | 461.0, 461.1, 461.2, 461.3, 461.8, 461.9, 462, 463, 464.00, 464.01, 464.10, 464.11, 464.20, 464.21, 464.30, 464.31, 464.4, 464.50, 464.51, 465.0, 465.8, 465.9, 481, 482.0, 482.1, 482.2, 482.30, 482.31, 482.32, 482.39, 482.40, 482.41, 482.42, 482.49, 482.81, 482.82, 482.83, 482.84, 482.89, 482.9, 485, | J01.00, J01.10, J01.20, J01.30, J01.40, J01.90, J02.9, J03.90, J04.0, J05.0, J04.10, J04.11, J04.2, J05.10, J05.11, J04.30, J04.31, J06.0, J06.9, J13, J18.1, J15.0, J15.1, J14, J15.4, J15.3, J15.20, J15.21, J15.211, J15.212, J15.29, J15.8, J15.5, J15.6, A48.1, J15.9, J18.0, J18.9, J44.1, J47.9, J47.1, J86.0, J86.9, J85.0, |

| | | |
|---|---|---|
| | 486, 491.21, 494.0, 494.1, 510.0, 510.9, 513.0, 513.1 | J85.1, J85.2, J85.3 |
| Digestive | 540.0, 540.1, 540.9, 541, 542, 562.01, 562.03, 562.11, 562.13, 566, 567.0, 567.1, 567.21, 567.22, 567.23, 567.29, 567.31, 567.38, 567.39, 567.81, 567.89, 567.9, 569.5, 569.83, 572.0, 572.1, 575.0, 575.10, 575.11, 575.12, 575.4, 575.5, 575.9 | K35.2, K35.3, K35.80, K35.89, K37, K36, K57.12, K57.13, K57.32, K57.33, K61.3, K61.1, K61.0, K67, K65.8, K65.0, K65.1, K65.2, K68.12, K68.19, K68.9, K65.3, K65.9, K63.0, K63.1, K75.0, K75.1, K81.0, K81.9, K81.1, K81.2, K82.2, K82.3, K82.9 |
| Genitourinary | 098.0, 098.10, 098.11, 098.12, 098.13, 098.14, 098.15, 098.16, 098.17, 098.19, 098.2, 098.30, 098.31, 098.32, 098.33, 098.34, 098.35, 098.36, 098.37, 098.39, 098.40, 098.41, 098.42, 098.43, 098.49, 098.50, 098.51, 098.52, 098.53, 098.59, 098.6, 098.7, 098.81, 098.82, 098.83, 098.84, 098.85, 098.86, 098.89, 590.00, 590.01, 590.10, 590.11, 590.2, 590.3, 590.80, 590.81, 590.9, 597.0, 601.0, 601.1, 601.2, 601.3, 601.4, 601.8, 601.9, 614.0, 614.1, 614.2, 614.3, 614.4, 614.5, 614.6, 614.7, 614.8, 614.9, 615.0, 615.1, 615.9, 616.0, 616.10, 616.11, 616.3, 616.4, 616.89, 616.9, 599.0 | A54.00, A54.29, A54.01, A54.22, A54.23, A54.03, A54.24, A54.21, A54.31, A54.32, A54.39, A54.33, A54.42, A54.49, A54.41, A54.40, A54.5, A54.6, A54.89, A54.81, A54.83, A54.85, A54.86, N11.0, N11.8, N10, N15.1, N28.84, N28.85, N28.86, N12, N16, N15.9, N34.0, N41.00, N41.01, N41.0, N41.10, N41.1, N41.11, N41.2, N41.3, N51, N41.8, N41.4, N41.9, N70.01, N70.03, N70.02, N70.13, N70.12, N70.11, N70.91, N70.93, N70.92, N73.0, N73.1, N73.2, N73.3, N73.6, N73.4, N73.8, N73.9, N71.0, N71.1, N71.9, N72, N76.3, N76.0, N76.2, N76.1, N77.1, N75.1, N76.4, N76.89, N75.9, N39.0 |
| Skin | 681.00, 681.01, 681.02, 681.10, 681.11, 681.9, 682.0, 682.1, 682.2, 682.3, 682.4, 682.5, 682.6, 682.7, 682.8, 682.9, 683, 686.00, 686.09, 686.1, 686.8, 686.9 | L03.029, L03.019, L03.039, L03.049, K12.2, L03.212, L03.211, L03.222, L03.221, L03.319, L03.329, L03.119, L03.129, L03.317, L03.818, L03.811, L03.898, L03.891, L03.91, L03.90, L04.9, L08.0, L08.89, L98.0, L08.9 |
| Musculoskeletal | 711.00, 711.01, 711.02, 711.03, 711.04, 711.05, 711.06, 711.07, 711.08, 711.09, 711.40, 711.41, 711.42, 711.43, 711.44, 711.45, 711.46, 711.47, 711.48, 711.49, 711.90, 711.91, 711.92, 711.93, 711.94, 711.95, 711.96, 711.97, 711.98, 711.99, 730.00, 730.01, 730.02, 730.03, 730.04, 730.05, 730.06, 730.07, 730.08, 730.09, 730.10, 730.11, 730.12, 730.13, 730.14, 730.15, 730.16, 730.17, 730.18, 730.19, 730.20, 730.21, 730.22, 730.23, 730.24, 730.25, 730.26, 730.27, 730.28, 730.29, 730.30, 730.31, 730.32, 730.33, 730.34, 730.35, 730.36, 730.37, 730.38, 730.39, 730.80, 730.81, 730.82, 730.83, 730.84, 730.85, 730.86, 730.87, 730.88, 730.89, 730.90, 730.91, 730.92, 730.93, 730.94, 730.95, 730.96, 730.97, 730.98, 730.99 | M00.9, M00.80, M00.00, M00.20, M00.10, M00.119, M00.819, M00.219, M00.019, M00.229, M00.829, M00.029, M00.129, M00.239, M00.039, M00.139, M00.839, M00.149, M00.249, M00.849, M00.049, M00.059, M00.159, M00.259, M00.859, M00.869, M00.069, M00.269, M00.169, M00.079, M00.279, M00.179, M00.879, M00.28, M00.18, M00.88, M00.08, M00.29, M00.09, M00.89, M00.19, M01.X0, M01.X19, M01.X29, M01.X39, M01.X49, M01.X59, M01.X69, M01.X79, M01.X8, M01.X9, M86.10, M86.20, M86.219, M86.119, M86.129, M86.229, M86.139, M86.239, M86.249, M86.149, M86.259, M86.159, M86.269, M86.169, |

| | | |
|---|---|---|
| | | M86.279, M86.179, M86.28, M86.18, M86.19, M86.29, M86.60, M86.619, M86.629, M86.639, M86.642, M86.659, M86.669, M86.679, M86.68, M86.69, M86.9, M46.20, M90.80, M90.819, M90.829, M90.839, M90.849, M90.859, M90.869, M90.879, M90.88, M90.89, M46.30 |
| Other | 790.7, 996.60, 996.61, 996.62, 996.63, 996.64, 996.65, 996.66, 996.67, 996.68, 996.69, 998.51, 998.59, 999.31, 999.32, 999.33, 999.34, 999.39 | R78.81, T85.79XA, T82.7XXA, T82.6XXA, T83.51XA, T83.6XXA, T83.59XA, T84.50XA, T84.7XXA, T84.60XA, T85.71XA, T81.4XXA, K68.11, T80.21XA, T80.219A, T80.211A, T80.212A, T80.22XA, T80.29XA, T88.0XXA |

*No codes for this category in this cohort.

**Supplementary File 1b. ICD-9-CM and ICD-10-CM codes for comorbidities.**

| | |
|---|---|
| **Congestive heart failure** | 428,428.1,428.2,428.21,428.22,428.23,428.3,428.31,428.32,428.33,428.4,428.41,428.42,428.43,428.9,I50.23,I50.22,I50.31,I50.9,I50.30,I50.814,I50.32,I50.21,I50.42,I50.20,I50.41,I50.40,I50.43,I50.33,I50.1,I50.84,I50.811,I50.83,I50.813,I50.810,I50.89,I50.82,I50.812 |
| **Chronic pulmonary disease** | 490,491,491.1,491.2,491.21,491.22,491.8,491.9,492,492.8,493,493.01,493.02,493.1,493.11,493.12,493.2,493.21,493.22,493.81,493.82,493.9,493.91,493.92,494,494.1,495,495.1,495.2,495.3,495.4,495.5,495.6,495.7,495.8,495.9,496,500,501,502,503,504,505,506.4,J40,J41.0,J41.1,J44.9,J44.1,J44.0,J41.8,J42,J43.9,J43.2,J43.1,J43.8,J43.0,J45.20,J45.40,J45.30,J45.50,J45.52,J45.32,J45.42,J45.22,J45.31,J45.41,J45.21,J45.51,J45.990,J45.991,J45.998,J45.909,J45.902,J45.901,J47.9,J47.1,J47.0,J67.0,J67.1,J67.2,J67.3,J67.4,J67.5,J67.6,J67.7,J67.8,J67.9,J60,J61,J62.8,J62.0,J63.0,J63.5,J63.3,J63.1,J63.2,J63.4,J63.6,J66.1,J66.2,J66.8,J66.0,J64,J65,J68.4 |
| **Cerebrovascular disease** | 430,431,432,432.1,432.9,433,433.01,433.1,433.11,433.2,433.21,433.3,433.31,433.8,433.81,433.9,433.91,434,434.01,434.1,434.11,434.9,434.91,435,435.1,435.2,435.3,435.8,435.9,436,437,437.1,437.2,437.3,437.4,437.5,437.6,437.7,437.8,437.9,438,438.1,438.11,438.12,438.13,438.14,438.19,438.2,438.21,438.22,438.3,438.31,438.32,438.4,438.41,438.42,438.5,438.51,438.52,438.53,438.6,438.7,438.81,438.82,438.83,438.84,438.85,438.89,438.9,I60.01,I60.31,I60.50,I60.02,I60.30,I60.4,I60.8,I60.52,I60.2,I60.7,I60.11,I60.51,I60.9,I60.32,I60.10,I60.12,I60.6,I60.00,I61.5,I61.6,I61.4,I61.9,I61.8,I61.3,I61.0,I61.2,I61.1,I62.1,I62.03,I62.01,I62.00,I62.02,I62.9,I65.1,I63.12,I63.02,I63.22,I65.22,I65.21,I65.29,I65.23,I63.032,I63.031,I63.132,I63.131,I63.139,I63.239,I63.033,I63.039,I63.231,I63.133,I63.233,I63.232,I65.09,I65.02,I65.01,I65.03,I63.212,I63.011,I63.213,I63.219,I63.211,I63.111,I63.013,I63.019,I63.119,I63.012,I63.113,I63.112,I65.8,I63.59,I63.19,I63.09,I65.9,I63.00,I63.20,I63.29,I63.10,I66.01,I66.23,I66.21,I66.12,I66.13,I66.02,I66.22,I66.09,I66.19,I66.03,I66.3,I66.11,I66.29,I63.39,I63.349,I63.311,I63.319,I63.343,I63.30,I63.312,I63.341,I63.332,I63.342,I63.333,I63.313,I63.323,I63.6,I63.329,I63.322,I63.331,I63.321,I63.339,I66.9,I63.439,I63.449,I63.40,I63.423,I63.429,I63.442,I63.422,I63.419,I63.433,I63.411,I63.421,I63.443,I63.49,I63.441,I63.431,I63.412,I63.432,I63.413,I66.8,I63.529,I63.9,I63.522,I63.539,I63.541,I63.533,I63.542,I63.531,I63.523,I63.50,I63.8,I63.543,I63.519,I63.521,I63.549,I63.512,I63.511,I63.532,I63.513,G45.0,G45.8,G46.1,G45.2,G46.2,G46.0,G45.1,I67.848,G45.9,I67.841,I67.89,I67.2,I67.81,I67.82,I67.4,I67.1,I67.7,I68.2,I67.5,I67.6,G45.4,G46.5,I68.0,G46.7,G46.3,G46.6,G46.8,G46.4,I68.8,I67.9,I69.311,I69.213,I69.010,I69.813,I69.012,I69.810,I69.118,I69.915,I69.315,I69.211,I69.815,I69.819,I69.913,I69.314,I69.019,I69.912,I69.219,I69.218,I69.011,I69.919,I69.914,I69.212,I69.013,I69.114,I69.313,I69.111,I69.214,I69.911,I69.811,I69.918,I69.310,I69.215,I69.115,I69.119,I69.018,I69.312,I69.210,I69.812,I69.818,I69.814,I69.910,I69.318,I69.014,I69.015,I69.112,I69.113,I69.110,I69.319,I69.928,I69.020,I69.320,I69.220,I69.120,I69.920,I69.820,I69.821,I69.921,I69.021,I69.121,I69.321,I69.221,I69.822,I69.222,I69.022,I69.922,I69.122,I69.322,I69.823,I69.223,I69.323,I69.123,I69.023,I69.923,I69.228,I69.328,I69.028,I69.828,I69.128,I69.059,I69.859,I69.359,I69.159,I69.959,I69.259,I69.951,I69.352,I69.051,I69.852,I69.052,I69.151,I69.851,I69.251,I69.152,I69.252,I69.351,I69.952,I69.054,I69.353,I69.154,I69.253,I69.954,I69.254,I |

| | |
|---|---|
| | 69.153,I69.053,I69.354,I69.953,I69.854,I69.853,I69.039,I69.839,I69.239,I69.139,I69.939,I69.339,I69.131,I69.932,I69.332,I69.931,I69.032,I69.832,I69.031,I69.232,I69.132,I69.331,I69.831,I69.231,I69.134,I69.334,I69.934,I69.234,I69.834,I69.333,I69.034,I69.033,I69.933,I69.133,I69.233,I69.833,I69.349,I69.849,I69.149,I69.049,I69.949,I69.249,I69.342,I69.142,I69.941,I69.041,I69.141,I69.942,I69.341,I69.241,I69.042,I69.842,I69.242,I69.841,I69.043,I69.344,I69.943,I69.343,I69.044,I69.844,I69.144,I69.244,I69.243,I69.944,I69.843,I69.143,I69.869,I69.069,I69.169,I69.969,I69.269,I69.369,I69.262,I69.061,I69.161,I69.861,I69.961,I69.362,I69.062,I69.162,I69.361,I69.962,I69.862,I69.261,I69.963,I69.163,I69.863,I69.864,I69.364,I69.164,I69.363,I69.064,I69.264,I69.063,I69.263,I69.964,I69.265,I69.365,I69.065,I69.165,I69.865,I69.965,I69.998,I69.890,I69.190,I69.090,I69.390,I69.290,I69.990,I69.891,I69.191,I69.291,I69.391,I69.991,I69.091,I69.092,I69.192,I69.292,I69.392,I69.892,I69.992,I69.893,I69.993,I69.093,I69.393,I69.193,I69.293,I69.398,I69.898,I69.298,I69.198,I69.098,I69.10,I69.80,I69.90,I69.30,I69.20,I69.00 |
| **Dementia** | 290,290.1,290.11,290.12,290.13,290.2,290.21,290.3,290.4,290.41,290.42,290.43,290.8,290.9,F03.90,F05,F01.50,F01.51 |
| **Diabetes with chronic complication*** | 250.4,250.41,250.42,250.43,250.5,250.51,250.52,250.53,250.6,250.61,250.62,250.63,E13.29,E11.21,E13.21,E13.22,E11.22,E11.29,E10.21,E10.29,E10.22,E11.65,E10.65,E13.3599,E13.3541,E11.3531,E13.3593,E13.319,E13.3521,E13.3543,E11.3532,E13.3542,E13.3311,E11.3513,E11.3212,E13.3539,E13.3519,E13.3513,E11.39,E13.3511,E13.3419,E13.3529,E13.3392,E11.3419,E13.3313,E13.3319,E11.3499,E11.3392,E13.3412,E13.3219,E11.3533,E11.3551,E11.3213,E13.36,E11.3491,E13.3299,E11.3393,E13.37X1,E11.3292,E11.3391,E13.3391,E11.3521,E11.3211,E11.3599,E11.3593,E13.3399,E11.3312,E11.3522,E11.3411,E11.3559,E13.3532,E13.37X3,E11.3523,E13.3291,E11.3493,E13.3523,E13.3552,E13.3591,E13.3553,E13.3213,E11.3542,E13.3411,E11.3399,E11.3539,E11.3592,E13.37X2,E11.37X1,E11.3511,E11.36,E11.37X3,E11.3553,E13.3292,E13.3413,E13.3312,E13.3551,E11.3549,E11.3413,E11.3519,E11.37X2,E11.3311,E13.3492,E11.3291,E13.39,E11.3529,E11.3591,E11.311,E13.3512,E11.3219,E13.3212,E13.3499,E11.3293,E13.3533,E11.319,E13.3493,E11.3299,E13.3592,E11.3541,E11.3412,E11.3552,E11.3543,E13.3393,E11.3319,E11.3313,E13.3211,E11.3512,E13.3559,E13.311,E13.3491,E13.37X9,E13.3549,E11.37X9,E13.3531,E13.3522,E13.3293,E11.3492,E10.3541,E10.3213,E10.3399,E10.3312,E10.3521,E10.3211,E10.3533,E10.37X3,E10.3551,E10.3492,E10.3219,E10.3522,E10.311,E10.3419,E10.3512,E10.3552,E10.36,E10.3593,E10.3413,E10.3319,E10.3313,E10.3513,E10.3212,E10.3542,E10.3499,E10.3599,E10.3539,E10.3549,E10.3559,E10.3393,E10.37X9,E10.3532,E10.3299,E10.3491,E10.3553,E10.3592,E10.3411,E10.3311,E10.3519,E10.3293,E10.3531,E10.3291,E10.3523,E10.3392,E10.3591,E10.3543,E10.3391,E10.3292,E10.3412,E10.3511,E10.319,E10.3493,E10.39,E10.37X2,E10.37X1,E10.3529,E11.44,E13.40,E11.610,E13.49,E11.40,E13.610,E11.41,E13.43,E13.42,E13.41,E11.42,E11.43,E11.49,E13.44,E10.49,E10.43,E10.44,E10.41,E10.42,E10.40,E10.610 |
| **Diabetes without chronic complication*** | 250,250.01,250.02,250.03,250.1,250.11,250.12,250.13,250.3,250.31,250.32,250.33,250.7,250.71,250.72,250.73,E13.9,E11.9,E10.9,E11.65,E10.65,E11.10,E13.10,E11.69,E10.10,E11.641,E13.641,E13.11,E11.11,E10.11,E10.641,E11.01,E11.59,E11.52,E13.59,E13.52,E11.51,E13.51,E10.52,E10.59,E10.51 |
| **Hemiplegia or paraplegia** | 344.1,343,343.1,343.2,343.3,343.4,343.8,343.9,G04.1,G82.22,G82.20,G82.21,G80.1,G80.2,G80.0,G80.8,G80.4,G80.9 |

| | |
|---|---|
| **AIDS/HIV** | 42,B20 |
| **Any malignancy, including lymphoma and leukemia, except malignant neoplasm of skin** | 140,140.1,140.3,140.4,140.5,140.6,140.8,140.9,141,141.1,141.2,141.3,141.4,141.5,141.6,141.8,141.9,142,142.1,142.2,142.8,142.9,143,143.1,143.8,143.9,144,144.1,144.8,144.9,145,145.1,145.2,145.3,145.4,145.5,145.6,145.8,145.9,146,146.1,146.2,146.3,146.4,146.5,146.6,146.7,146.8,146.9,147,147.1,147.2,147.3,147.8,147.9,148,148.1,148.2,148.3,148.8,148.9,149,149.1,149.8,149.9,150,150.1,150.2,150.3,150.4,150.5,150.8,150.9,151,151.1,151.2,151.3,151.4,151.5,151.6,151.8,151.9,152,152.1,152.2,152.3,152.8,152.9,153,153.1,153.2,153.3,153.4,153.5,153.6,153.7,153.8,153.9,154,154.1,154.2,154.3,154.8,155,155.1,155.2,156,156.1,156.2,156.8,156.9,157,157.1,157.2,157.3,157.4,157.8,157.9,158,158.8,158.9,159,159.1,159.8,159.9,160,160.1,160.2,160.3,160.4,160.5,160.8,160.9,161,161.1,161.2,161.3,161.8,161.9,162,162.2,162.3,162.4,162.5,162.8,162.9,163,163.1,163.8,163.9,164,164.1,164.2,164.3,164.8,164.9,165,165.8,165.9,170,170.1,170.2,170.3,170.4,170.5,170.6,170.7,170.8,170.9,171,171.2,171.3,171.4,171.5,171.6,171.7,171.8,171.9,172,172.1,172.2,172.3,172.4,172.5,172.6,172.7,172.8,172.9,174,174.1,174.2,174.3,174.4,174.5,174.6,174.8,174.9,175,175.9,176,176.1,176.2,176.3,176.4,176.5,176.8,176.9,179,180,180.1,180.8,180.9,181,182,182.1,182.8,183,183.2,183.3,183.4,183.5,183.8,183.9,184,184.1,184.2,184.3,184.4,184.8,184.9,185,186,186.9,187.1,187.2,187.3,187.4,187.5,187.6,187.7,187.8,187.9,188,188.1,188.2,188.3,188.4,188.5,188.6,188.7,188.8,188.9,189,189.1,189.2,189.3,189.4,189.8,189.9,190,190.1,190.2,190.3,190.4,190.5,190.6,190.7,190.8,190.9,191,191.1,191.2,191.3,191.4,191.5,191.6,191.7,191.8,191.9,192,192.1,192.2,192.3,192.8,192.9,193,194,194.1,194.3,194.4,194.5,194.6,194.8,194.9,195,195.1,195.2,195.3,195.4,195.5,195.8,200,200.01,200.02,200.03,200.04,200.05,200.06,200.07,200.08,200.1,200.11,200.12,200.13,200.14,200.15,200.16,200.17,200.18,200.2,200.21,200.22,200.23,200.24,200.25,200.26,200.27,200.28,200.3,200.31,200.32,200.33,200.34,200.35,200.36,200.37,200.38,200.4,200.41,200.42,200.43,200.44,200.45,200.46,200.47,200.48,200.5,200.51,200.52,200.53,200.54,200.55,200.56,200.57,200.58,200.6,200.61,200.62,200.63,200.64,200.65,200.66,200.67,200.68,200.7,200.71,200.72,200.73,200.74,200.75,200.76,200.77,200.78,200.8,200.81,200.82,200.83,200.84,200.85,200.86,200.87,200.88,201,201.01,201.02,201.03,201.04,201.05,201.06,201.07,201.08,201.1,201.11,201.12,201.13,201.14,201.15,201.16,201.17,201.18,201.2,201.21,201.22,201.23,201.24,201.25,201.26,201.27,201.28,201.4,201.41,201.42,201.43,201.44,201.45,201.46,201.47,201.48,201.5,201.51,201.52,201.53,201.54,201.55,201.56,201.57,201.58,201.6,201.61,201.62,201.63,201.64,201.65,201.66,201.67,201.68,201.7,201.71,201.72,201.73,201.74,201.75,201.76,201.77,201.78,201.9,201.91,201.92,201.93,201.94,201.95,201.96,201.97,201.98,202,202.01,202.02,202.03,202.04,202.05,202.06,202.07,202.08,202.1,202.11,202.12,202.13,202.14,202.15,202.16,202.17,202.18,202.2,202.21,202.22,202.23,202.24,202.25,202.26,202.27,202.28,202.3,202.31,202.32,202.33,202.34,202.35,202.36,202.37,202.38,202.4,202.41,202.42,202.43,202.44,202.45,202.46,202.47,202.48,202.5,202.51,202.52,202.53,202.54,202.55,202.56,202.57,202.58,202.6,202.61,202.62,202.63,202.64,202.65,202.66,202.67,202.68,202.7,202.71,202.72,202.73,202.74,202.75,202.76,202.77,202.78,202.8,202.81,202.82,202.83,202.84,202.85,202.86,202.87,202.88,202.9,202.91,202.92,202.93,202.94,202.95,202.96,202.97,202.98,203,203.01,203.02,203.1,203.11,203.12,203.8,203.81,203.82,204,204.01,204.02,204.1,204.11,204.12,204.2,204.21,204.22,204.8,204.81,204.82,204.9,204.91,204.92,205,2 |

| | |
|---|---|
| | 05.01,205.02,205.1,205.11,205.12,205.2,205.21,205.22,205.3,205.31,205.32,205.8,205.81,205.82,205.9,205.91,205.92,206,206.01,206.02,206.1,206.11,206.12,206.2,206.21,206.22,206.8,206.81,206.82,206.9,206.91,206.92,207,207.01,207.02,207.1,207.11,207.12,207.2,207.21,207.22,207.8,207.81,207.82,208,208.01,208.02,208.1,208.11,208.12,208.2,208.21,208.22,208.8,208.81,208.82,208.9,208.91,208.92,C00.0,C00.1,C00.3,C00.4,C00.5,C00.6,C00.8,C00.9,C00.2,C01,C02.0,C02.1,C02.2,C02.3,C02.8,C02.4,C02.9,C07,C08.0,C08.1,C08.9,C03.0,C03.1,C03.9,C04.0,C04.1,C04.8,C04.9,C06.0,C06.1,C05.0,C05.1,C05.2,C05.9,C05.8,C06.2,C06.89,C06.80,C06.9,C09.9,C09.8,C09.0,C09.1,C10.0,C10.1,C10.8,C10.2,C10.3,C10.4,C10.9,C11.0,C11.1,C11.2,C11.3,C11.8,C11.9,C13.0,C12,C13.1,C13.2,C13.8,C13.9,C14.0,C14.2,C14.8,C15.3,C15.4,C15.5,C15.8,C15.9,C16.0,C16.4,C16.3,C16.1,C16.2,C16.5,C16.6,C16.8,C16.9,C17.0,C17.1,C17.2,C17.3,C17.8,C17.9,C18.3,C18.4,C18.6,C18.7,C18.0,C18.1,C18.2,C18.5,C18.8,C18.9,C19,C20,C21.1,C21.0,C21.8,C21.2,C22.3,C22.7,C22.8,C22.4,C22.2,C22.0,C22.1,C22.9,C23,C24.0,C24.1,C24.8,C24.9,C25.0,C25.1,C25.2,C25.3,C25.4,C25.8,C25.7,C25.9,C48.0,C45.1,C48.8,C48.1,C48.2,C26.0,C26.1,C26.9,C30.0,C30.1,C31.0,C31.1,C31.2,C31.3,C31.8,C31.9,C32.0,C32.1,C32.2,C32.3,C32.8,C32.9,C33,C34.00,C34.02,C34.01,C34.12,C34.10,C34.11,C34.2,C34.30,C34.32,C34.31,C34.82,C34.80,C34.81,C34.91,C34.90,C34.92,C38.4,C45.0,C37,C38.0,C45.2,C38.1,C38.2,C38.8,C38.3,C39.0,C39.9,C41.0,C41.1,C41.2,C41.3,C40.01,C40.00,C40.02,C40.12,C40.11,C40.10,C41.4,C40.20,C40.22,C40.21,C40.30,C40.32,C40.31,C40.92,C40.82,C40.80,C40.81,C40.90,C41.9,C40.91,C49.0,C47.0,C49.10,C47.12,C47.11,C47.10,C49.11,C49.12,C49.22,C47.21,C47.22,C47.20,C49.21,C49.20,C47.3,C49.3,C49.A1,C49.A2,C49.A4,C49.4,C49.A3,C49.A5,C49.A0,C49.A9,C47.4,C47.5,C49.5,C49.6,C47.6,C49.8,C47.8,C47.9,C49.9,D03.0,C43.0,D03.11,C43.10,C43.12,D03.10,D03.12,C43.11,C43.22,C43.20,D03.20,D03.22,C43.21,D03.21,C43.30,C43.31,C43.39,D03.39,D03.30,C43.4,D03.4,D03.59,C43.52,D03.51,D03.52,C43.59,C43.51,D03.61,C43.60,C43.61,D03.62,C43.62,D03.60,C43.72,D03.72,C43.71,C43.70,D03.71,D03.70,D03.8,C43.8,C43.9,D03.9,C50.011,C50.019,C50.012,C50.119,C50.112,C50.111,C50.212,C50.211,C50.219,C50.311,C50.319,C50.312,C50.419,C50.411,C50.412,C50.511,C50.519,C50.512,C50.612,C50.619,C50.611,C50.812,C50.811,C50.819,C50.911,C50.912,C50.919,C50.022,C50.021,C50.029,C50.122,C50.221,C50.229,C50.529,C50.329,C50.629,C50.829,C50.621,C50.121,C50.921,C50.929,C50.821,C50.822,C50.622,C50.922,C50.421,C50.321,C50.521,C50.429,C50.322,C50.522,C50.222,C50.129,C50.422,C46.0,C46.1,C46.2,C46.4,C46.51,C46.50,C46.52,C46.3,C46.7,C46.9,C55,C53.0,C53.1,C53.8,C53.9,C58,C54.9,C54.3,C54.1,C54.2,C54.0,C54.8,C56.2,C56.9,C56.1,C57.01,C57.00,C57.02,C57.10,C57.12,C57.11,C57.3,C57.20,C57.21,C57.22,C57.4,C52,C51.0,C51.1,C51.2,C51.9,C57.7,C51.8,C57.8,C57.9,C61,C62.01,C62.02,C62.00,C62.90,C62.12,C62.92,C62.11,C62.91,C62.10,C60.0,C60.1,C60.2,C60.9,C63.02,C63.01,C63.00,C63.12,C63.10,C63.11,C63.2,C60.8,C63.7,C63.8,C63.9,C67.0,C67.1,C67.2,C67.3,C67.4,C67.5,C67.6,C67.7,C67.8,C67.9,C64.1,C64.9,C64.2,C65.2,C65.1,C65.9,C66.9,C66.2,C66.1,C68.0,C68.1,C68.8,C68.9,C69.40,C69.41,C69.42,C69.62,C69.60,C69.61,C69.51,C69.52,C69.50,C69.01,C69.00,C69.02,C69.10,C69.12,C69.11,C69.21,C69.20,C69.22,C69.31,C69.30,C69.32,C69.80,C69.82,C69.81,C69.92,C69.91,C69.90,C71.0,C71.1,C71.2,C71.3,C71.4,C71.5,C71.6,C71.7,C71.8,C71.9,C72.20,C72.30,C72.59,C72.21,C72.40,C72.22,C72.50,C72.31,C72.32,C72.42,C72.41,C70.9,C70.0,C72.1,C72.0,C70.1,C72.9,C73,C74.90,C74.10,C74.12,C74.11,C74.92,C74.02,C74.01,C74.91,C74.00,C75.0,C75.1,C75.2,C75.3,C75.4,C75.5,C75.8,C75.9,C |

| | |
|---|---|
| | 76.0,C76.1,C76.2,C76.3,C76.42,C76.40,C76.41,C76.50,C76.52,C76.51,C76.8,C45.7,C83.39,C83.30,C83.31,C83.32,C83.33,C83.34,C83.35,C83.36,C83.37,C83.38,C83.50,C83.59,C83.51,C83.52,C83.53,C83.54,C83.55,C83.56,C83.57,C83.58,C83.70,C83.79,C83.71,C83.72,C83.73,C83.74,C83.75,C83.76,C83.77,C83.78,C83.80,C83.89,C88.4,C83.81,C83.82,C83.83,C83.84,C83.85,C83.86,C83.87,C83.88,C83.10,C83.19,C83.11,C83.12,C83.13,C83.14,C83.15,C83.16,C83.17,C83.18,C84.79,C84.60,C84.69,C84.70,C84.71,C84.61,C84.62,C84.72,C84.73,C84.63,C84.74,C84.64,C84.75,C84.65,C84.76,C84.66,C84.67,C84.77,C84.78,C84.68,C85.20,C85.29,C85.21,C85.22,C85.23,C85.24,C85.25,C85.26,C85.27,C85.28,C83.99,C83.09,C86.5,C86.6,C83.90,C83.00,C83.01,C83.91,C83.92,C83.02,C83.03,C83.93,C83.94,C83.04,C83.05,C83.95,C83.06,C83.96,C83.07,C83.97,C83.98,C83.08,C81.70,C81.79,C81.71,C81.72,C81.73,C81.74,C81.75,C81.76,C81.77,C81.78,C81.49,C81.00,C81.09,C81.40,C81.41,C81.01,C81.42,C81.02,C81.43,C81.03,C81.44,C81.04,C81.45,C81.05,C81.46,C81.06,C81.47,C81.07,C81.48,C81.08,C81.19,C81.10,C81.11,C81.12,C81.13,C81.14,C81.15,C81.16,C81.17,C81.18,C81.29,C81.20,C81.21,C81.22,C81.23,C81.24,C81.25,C81.26,C81.27,C81.28,C81.39,C81.30,C81.31,C81.32,C81.33,C81.34,C81.35,C81.36,C81.37,C81.38,C81.90,C81.99,C81.91,C81.92,C81.93,C81.94,C81.95,C81.96,C81.97,C81.98,C82.29,C82.49,C82.40,C82.09,C82.69,C82.39,C82.20,C82.80,C82.60,C82.30,C82.19,C82.10,C82.99,C82.00,C82.90,C82.89,C82.81,C82.61,C82.21,C82.91,C82.11,C82.01,C82.41,C82.31,C82.22,C82.62,C82.02,C82.32,C82.82,C82.42,C82.92,C82.12,C82.43,C82.83,C82.03,C82.33,C82.13,C82.63,C82.93,C82.23,C82.94,C82.14,C82.64,C82.24,C82.84,C82.34,C82.44,C82.04,C82.05,C82.15,C82.85,C82.35,C82.25,C82.45,C82.95,C82.65,C82.06,C82.36,C82.86,C82.46,C82.96,C82.66,C82.26,C82.16,C82.47,C82.67,C82.97,C82.07,C82.87,C82.37,C82.27,C82.17,C82.98,C82.18,C82.88,C82.28,C82.38,C82.08,C82.68,C82.48,C84.09,C84.00,C84.01,C84.02,C84.03,C84.04,C84.05,C84.06,C84.07,C84.08,C84.19,C84.10,C84.11,C84.12,C84.13,C84.14,C84.15,C84.16,C84.17,C84.18,C96.A,C91.42,C91.40,C91.41,C96.0,C96.29,C96.21,C96.20,C96.22,C84.40,C84.49,C84.41,C84.42,C84.43,C84.44,C84.45,C84.46,C84.47,C84.48,C85.80,C86.4,C84.Z9,C84.A9,C85.19,C84.Z0,C85.99,C84.A0,C82.50,C82.59,C85.90,C84.90,C84.99,C85.89,C85.10,C82.51,C84.91,C85.91,C84.Z1,C84.A1,C86.0,C85.11,C85.81,C84.92,C84.A2,C82.52,C85.12,C84.Z2,C85.82,C85.92,C85.13,C84.Z3,C82.53,C84.93,C86.3,C84.A3,C85.93,C86.2,C85.83,C85.14,C85.84,C84.Z4,C82.54,C85.94,C84.A4,C84.94,C84.A5,C85.85,C82.55,C84.95,C85.15,C84.Z5,C85.95,C85.86,C85.96,C84.A6,C84.96,C82.56,C85.16,C84.Z6,C85.17,C84.Z7,C85.87,C82.57,C85.97,C86.1,C84.97,C84.A7,C85.18,C84.Z8,C84.98,C85.98,C84.A8,C82.58,C85.88,C96.4,C96.9,C96.Z,C90.00,C90.01,C90.02,C90.10,C90.11,C90.12,C88.8,C88.3,C90.20,C88.9,C88.2,C90.30,C90.31,C90.21,C90.32,C90.22,C91.00,C91.01,C91.02,C91.10,C91.11,C91.12,C91.Z0,C91.Z1,C91.Z2,C91.50,C91.30,C91.A0,C91.60,C91.61,C91.31,C91.51,C91.A1,C91.52,C91.32,C91.A2,C91.62,C91.90,C91.91,C91.92,C92.60,C92.50,C92.00,C92.40,C92.A0,C92.41,C92.51,C92.01,C92.61,C92.A1,C92.42,C92.02,C92.62,C92.52,C92.A2,C92.10,C92.11,C92.12,C92.20,C92.21,C92.22,C92.30,C92.31,C92.32,C92.Z0,C92.Z1,C92.Z2,C92.90,C92.91,C92.92,C93.00,C93.01,C93.02,C93.10,C93.11,C93.12,C93.90,C93.91,C93.92,C93.30,C93.Z0,C93.31,C93.Z1,C93.Z2,C93.32,C94.00,C94.01,C94.02,D45,C94.20,C94.21,C94.22,C94.80,C94.30,C94.31,C94.81,C94.32,C94.82,C95.00,C95.01,C95.02,C95.10,C95.11,C95.12,C95.90,C95.91,C95.92 |

| | |
|---|---|
| **Myocardial infarction** | 410,410.01,410.02,410.1,410.11,410.12,410.2,410.21,410.22,410.3,410.31,410.32,410.4,410.41,410.42,410.5,410.51,410.52,410.6,410.61,410.62,410.7,410.71,410.72,410.8,410.81,410.82,410.9,410.91,410.92,412,I21.09,I22.0,I21.02,I21.01,I21.19,I22.1,I21.11,I21.29,I22.8,I21.4,I22.2,I21.21,I21.3,I21.A9,I21.9,I21.A1,I22.9,I25.2 |
| **Mild liver disease*** | 571.2,571.4,571.41,571.42,571.49,571.5,571.6,K70.30,K70.2,K70.31,K73.9,K73.0,K75.4,K73.8,K73.1,K73.2,K74.0,K74.60,K74.69,K74.4,K74.3,K74.5 |
| **Moderate or severe liver disease*** | 456,456.1,456.2,456.21,572.2,572.3,572.4,572.8,I85.01,I85.00,I85.11,I85.10,K72.11,K70.41,K72.91,K72.90,K72.01,K71.11,K76.6,K76.7,K72.10 |
| **Peptic ulcer disease** | 531,531.01,531.1,531.11,531.2,531.21,531.3,531.31,531.4,531.41,531.5,531.51,531.6,531.61,531.7,531.71,531.9,531.91,532,532.01,532.1,532.11,532.2,532.21,532.3,532.31,532.4,532.41,532.5,532.51,532.6,532.61,532.7,532.71,532.9,532.91,533,533.01,533.1,533.11,533.2,533.21,533.3,533.31,533.4,533.41,533.5,533.51,533.6,533.61,533.7,533.71,533.9,533.91,534,534.01,534.1,534.11,534.2,534.21,534.3,534.31,534.4,534.41,534.5,534.51,534.6,534.61,534.7,534.71,534.9,534.91,K25.0,K56.699,K25.1,K25.2,K25.3,K25.4,K25.5,K25.6,K25.7,K25.9,K26.0,K26.1,K26.2,K26.3,K26.4,K26.5,K26.6,K26.7,K26.9,K27.0,K27.1,K27.2,K27.3,K27.4,K27.5,K27.6,K27.7,K27.9,K28.0,K28.1,K28.2,K28.3,K28.4,K28.5,K28.6,K28.7,K28.9 |
| **Peripheral vascular disease** | 443.9,441,441.01,441.02,441.03,441.1,441.2,441.3,441.4,441.5,441.6,441.7,441.9,785.4,V43.4,I73.9,I71.00,I71.01,I71.02,I71.03,I71.1,I71.2,I71.3,I71.4,I71.8,I71.5,I71.6,I79.0,I71.9,I70.663,I70.563,I70.661,E09.52,I70.761,I70.763,I70.369,I70.769,E08.52,E13.52,I70.363,I70.469,I70.562,I73.01,I70.461,E11.52,I70.462,I70.762,E10.52,I70.768,I70.361,I70.561,I70.668,I70.368,I70.569,I70.568,I70.662,I70.669,I70.362,I96,I70.463,I70.468,Z95.828,Z95.820,38.48,04R.Y47Z,04R.M4JZ,04R.Q07Z,04R.U4JZ,04R.S4JZ,04R.Q0JZ,04R.U07Z,04R.R0JZ,04R.Q47Z,04R.N47Z,04R.T0JZ,04R.N0KZ,04R.L4KZ,04R.V47Z,04R.K47Z,04R.Q0KZ,04R.K4JZ,04R.Y07Z,04R.W0KZ,04R.V0JZ,04R.M47Z,04R.P47Z,04R.T07Z,04R.M4KZ,04R.S0KZ,04R.P07Z,04R.U0KZ,04R.P0KZ,04R.L07Z,04R.K0KZ,04R.R07Z,04R.Y0KZ,04R.K0JZ,04R.Y0JZ,04R.N07Z,04R.R4JZ,04R.U0JZ,04R.W4JZ,04R.L4JZ,04R.T4KZ,04R.R0KZ,04R.M0JZ,04R.U4KZ,04R.K07Z,04R.S47Z,04R.S0JZ,04R.V0KZ,04R.V07Z,04R.R4KZ,04R.U47Z,04R.N4JZ,04R.W4KZ,04R.P4JZ,04R.N4KZ,04R.Q4KZ,04R.K4KZ,04R.Y4KZ,04R.N0JZ,04R.V4KZ,04R.L0KZ,04R.Q4JZ,04R.W0JZ,04R.V4JZ,04R.S07Z,04R.W47Z,04R.T47Z,04R.T0KZ,04R.L47Z,04R.S4KZ,04R.M0KZ,04R.T4JZ,04R.W07Z,04R.R47Z,04R.P4KZ,04R.Y4JZ,04R.P0JZ,04R.M07Z,04R.L0JZ |
| **Renal disease** | 582,582.0,582.1,582.2,582.4,582.81,582.89,582.9,583,583.0,583.1,583.2,583.4,583.6,583.7,585.1,585.2,585.3,585.4,585.5,585.6,585.9,586,588,588.0,588.1,588.81,588.89,588.9,N03.2,N03.1,N03.3,N03.4,N03.7,N03.6,N03.5,N03.8,N08,N03.0,N03.9,N05.9,N05.2,N06.2,N07.2,N05.5,N05.3,N07.5,N06.3,N07.4,N06.5,N05.4,N06.4,N07.3,N17.1,N17.2,N18.1,N18.2,N18.3,N18.4,N18.5,N18.6,N18.9,N19,N25.0,N25.1,N25.81,N25.89,N25.9 |
| **Rheumatic disease** | 710,710.1,710.4,714,714.1,714.2,714.81,725,M32.8,M32.19,M32.14,M32.13,M32.12,M32.0,M32.10,M32.15,M32.9,M32.11,M34.1,M34.81,M34.2,M34.9,M34.83,M34.0,M34.89,M34.82,M33.22,M33.20,M33.21,M33.29,M06.249,M05.872,M05.549,M05.451,M06.071,M05.459,M05.461,M06.311,M06.821,M06.849,M05.59,M05.529,M05.452,M05.449,M06.339,M05.419,M05.442,M05.412,M05.71 |

| | |
|---|---|
| | 2,M05.50,M05.571,M05.552,M05.469,M06.859,M06.841,M05.852,M06.212,M05.779,M06.352,M06.251,M05.749,M05.579,M05.811,M05.831,M05.462,M05.862,M06.00,M05.721,M05.541,M05.522,M06.28,M05.869,M05.421,M06.219,M06.279,M05.722,M06.871,M05.732,M06.869,M05.742,M06.862,M06.839,M06.341,M05.812,M06.88,M06.831,M06.372,M05.819,M06.362,M05.471,M05.849,M05.851,M05.859,M06.051,M06.332,M06.271,M06.261,M06.259,M05.871,M06.241,M06.239,M06.231,M06.229,M06.022,M05.752,M06.851,M06.012,M06.371,M06.042,M05.9,M06.262,M05.79,M05.829,M06.069,M05.821,M06.312,M06.321,M05.561,M06.019,M06.322,M05.711,M05.432,M06.252,M06.029,M05.80,M05.739,M05.842,M06.329,M06.822,M06.842,M06.021,M06.89,M05.839,M06.811,M06.361,M05.731,M05.719,M06.379,M06.879,M05.772,M06.039,M06.072,M06.812,M05.521,M06.30,M06.29,M06.031,M05.551,M06.351,M05.572,M06.062,M06.061,M06.221,M06.359,M05.441,M05.769,M06.852,M05.472,M06.272,M05.40,M05.569,M06.269,M06.832,M06.059,M06.222,M05.841,M06.011,M05.89,M06.319,M05.771,M05.512,M05.70,M06.211,M05.422,M05.532,M05.411,M05.429,M05.562,M05.531,M05.49,M06.242,M06.08,M05.559,M05.861,M05.431,M06.369,M06.38,M06.052,M05.822,M06.349,M06.872,M05.832,M05.511,M06.9,M05.479,M06.032,M06.39,M05.539,M06.041,M05.879,M06.819,M05.519,M06.079,M06.20,M05.751,M05.741,M05.762,M06.331,M06.80,M06.342,M05.439,M06.09,M05.729,M06.232,M05.761,M06.861,M06.829,M05.759,M05.542,M06.049,M05.071,M05.041,M05.019,M05.069,M05.051,M05.00,M05.079,M05.072,M05.021,M05.061,M05.032,M05.029,M05.022,M05.042,M05.012,M05.062,M05.052,M05.09,M05.059,M05.039,M05.049,M05.031,M05.011,M05.271,M05.319,M05.279,M05.651,M05.219,M05.361,M05.679,M05.622,M05.659,M05.652,M05.639,M05.372,M05.212,M05.29,M05.262,M05.379,M05.312,M05.231,M05.222,M05.341,M05.642,M05.311,M05.362,M05.252,M05.672,M05.631,M05.661,M05.352,M05.229,M05.332,M05.69,M05.359,M05.351,M05.272,M05.249,M05.211,M05.611,M05.331,M05.669,M05.339,M05.621,M05.261,M05.322,M05.269,M05.20,M05.251,M05.371,M05.39,M05.641,M05.259,M05.241,M05.612,M05.662,M06.1,M05.321,M05.632,M05.619,M05.329,M05.239,M05.342,M05.369,M05.629,M05.30,M05.221,M05.349,M05.649,M05.671,M05.242,M05.232,M05.60,M05.171,M05.162,M05.122,M05.132,M05.149,M05.141,M05.172,M05.179,M05.161,M05.111,M05.119,M05.10,M05.129,M05.142,M05.151,M05.169,M05.159,M05.19,M05.112,M05.121,M05.139,M05.131,M05.152,M35.3 |
| **Metastatic solid tumor** | 196,196.1,196.2,196.3,196.5,196.6,196.8,196.9,197,197.1,197.2,197.3,197.4,197.5,197.6,197.7,197.8,198,198.1,198.2,198.3,198.4,198.5,198.6,198.7,198.81,198.82,198.89,199,199.1,C77.0,C77.1,C77.2,C77.3,C77.4,C77.5,C77.8,C77.9,C78.00,C78.01,C78.02,C78.1,C78.2,C78.30,C78.39,C78.4,C78.5,C78.6,C78.7,C78.89,C78.80,C79.01,C79.00,C79.02,C79.11,C79.10,C79.19,C79.2,C79.31,C79.49,C79.40,C79.32,C79.52,C79.51,C79.61,C79.62,C79.60,C79.70,C79.71,C79.72,C79.81,C79.82,C79.89,C79.9,C80.0,C80.1,C45.9 |

*****The categories of diabetes with and without complications were treated as two mutually exclusive groups, as were the categories of mild and moderate to severe liver disease.

**Supplementary File 1c. List of codes to identify severe renal diseases.**

| | |
|---|---|
| N18.4 | Chronic kidney disease, stage 4 (severe) |
| N18.5 | Chronic kidney disease, stage 5 |
| N18.6 | End stage renal disease |
| N18.9 | Chronic kidney disease, unspecified |
| 585.4 | Chronic kidney disease, Stage IV (severe) |
| 585.5 | Chronic kidney disease, Stage V |
| 585.6 | End stage renal disease |
| 585.9 | Chronic kidney disease, unspecified |
| 586 | Renal failure, unspecified |
| Z99.2 | Dependence on renal dialysis |
| Z49.0 | Preparatory care for renal dialysis. |
| Z49.31 | Encounter for adequacy testing for hemodialysis |
| 39.95 | Hemodialysis |
| V45.11 | Renal dialysis status |
| V56.0 | Encounter for extracorporeal dialysis |

**Supplementary File 1d. Association between *APOL1* high-risk genotypes and sepsis-related outcomes**

| Outcome | *APOL1* low-risk (n=1,881) | *APOL1* high-risk (n=361) | Odds Ratio | 95% CI | p-value |
|---|---|---|---|---|---|
| | n events (%) | n events (%) | | | |
| Sepsis | 460 (24.5) | 105 (29.1) | 1.29 | 1.00-1.67 | 0.047 |
| Septic shock | 131 (7.0) | 32 (8.9) | 1.30 | 0.86-1.95 | 0.211 |
| Renal failure | 236 (12.5) | 67 (18.6) | 1.64 | 1.21-2.22 | 0.001 |
| Respiratory failure | 122 (6.5) | 14 (3.9) | 0.57 | 0.33-1.01 | 0.055 |
| Hematologic failure | 87 (4.6) | 15 (4.2) | 0.86 | 0.49-1.51 | 0.600 |
| Cardiovascular failure | 77 (4.1) | 14 (3.9) | 0.89 | 0.50-1.60 | 0.698 |
| Hepatic failure | 70 (3.7) | 13 (3.6) | 1.02 | 0.56-1.88 | 0.941 |
| Short-term mortality | 74 (3.9) | 10 (2.8) | 0.76 | 0.32-1.81 | 0.315 |

**Supplementary File 1e. Demographic summary of patients in the PheWAS analysis.**

| | | Total (n=14,713) | *APOL1* low-risk (n=12,625) | *APOL1* high-risk (n=2,088) | p-value |
|---|---|---|---|---|---|
| Gender | Female, n (%) | 1269 (60.8) | 9063 (61.6) | 7794 (61.7) | 0.42 |
| | Male, n (%) | 819 (39.2) | 5650 (38.4) | 4831 (38.3) | |
| Age (at recent visit), median (Q1-Q3) | | 44 (27-61) | 44 (27-61) | 46 (30-61) | 0.001 |
| EHR length (years), median (Q1-Q3) | | 10.7 (4.9-18.1) | 10.7 (4.8-18.0) | 10.8 (5.2-18.5) | 0.13 |
